# It Takes 52 to Recruit One: Recruitment Barriers in Mechanistic Stroke Neurorehabilitation

**DOI:** 10.64898/2026.08.24.26361192

**Authors:** Anne G. Gerding, Christiane M. Thiel

**Affiliations:** Biological Psychology Lab, Department of Psychology, School of Medicine and Health Sciences, Carl von Ossietzky Universität Oldenburg, Oldenburg, Germany; Rehabilitationszentrum Oldenburg, Klinik für Neurologie, Oldenburg, Germany; Research Center Neurosensory Science, Carl von Ossietzky Universität Oldenburg, Oldenburg, Germany; Cluster of Excellence “Hearing4all”, Carl von Ossietzky Universität Oldenburg, Oldenburg, Germany

**Keywords:** Stroke, Patient Recruitment, Neurorehabilitation, Eligibility Criteria, Patient Selection, Recruitment Efficiency, Screening-to-enrollment Ratio

## Abstract

**BACKGROUND:** Recruitment in stroke neurorehabilitation trials is often difficult, particularly in studies requiring MRI and repeated laboratory visits. The recruitment efficiency was analyzed to identify the major barriers to enrollment in a stroke neurorehabilitation trial.

**METHODS:** In this observational screening study, 1201 patients were screened at a neurological rehabilitation center in Germany between October 2023 and February 2026. Recruitment barriers were analyzed using a stepwise recruitment flow approach.

**RESULTS:** Of 678 patients with ischemic stroke, 13 were ultimately enrolled (1.9%; 1.1% of all 1201 screened rehabilitation patients). The most common exclusion reasons were strict clinical eligibility criteria (52.2%), travel distance to the study center (23.9%), and predefined age restrictions (17.9%). Recruitment losses occurred across multiple stages of the screening process.

**CONCLUSION:** Recruitment in stroke neurorehabilitation trials is strongly limited by restrictive study criteria and logistical barriers. More pragmatic and inclusive study designs may improve recruitment efficiency and better reflect real-world stroke populations

## 1 Introduction

Successful recruitment is a critical determinant of success in clinical rehabilitation research. Insufficient recruitment can lead to underpowered studies, prolonged study duration or increased costs. In stroke rehabilitation research, these challenges are particularly pronounced due to the complex clinical profiles of patients, strict eligibility criteria, and substantial logistical demands associated with participation (1).

Previous studies have consistently demonstrated low recruitment efficiency in stroke rehabilitation and neuromodulation trials. Reported inclusion rates are often extremely low, with only approximately 1–8% of screened patients ultimately enrolled in clinical studies. Ferreira et al. highlighted that highly restrictive eligibility criteria in stroke neurorecovery trials systematically exclude large proportions of the real-world stroke population (2). Similarly, Potter-Baker et al. reported that non-invasive brain stimulation studies frequently enroll only a small and highly selected subgroup of stroke survivors (3). Hodics et al. further demonstrated that clinical, logistical, and socioeconomic barriers substantially reduce enrollment, particularly in diverse patient populations (4). Systematic evidence provided by McGill et al. additionally showed that recruitment in stroke rehabilitation randomized controlled trials is generally slow and inefficient across settings (1).

Existing literature about recruitment efficiency in stroke rehabilitation has primarily focused on individual recruitment barriers or overall recruitment rates and identified a range of clinical, demographic, logistical, and organizational barriers to study participation. Previous studies have highlighted factors such as stroke severity, comorbidities, MRI contraindications, cognitive and functional impairment, transportation difficulties, age and socioeconomic status as important determinants of recruitment success (5,6). In addition, recruitment efficiency appears to vary according to the clinical setting and stage of stroke recovery.

Systematic evidence suggests differences in recruitment between hospital- and home-based settings and between acute and later stages after stroke (1). McGill et al. (2020) showed that recruitment of acute stroke survivors within a hospital setting has been highlighted as a problematic recruitment area and most patients (38%) were recruited in the chronic phase (>6 months post stroke). However, the optimal stage and setting for recruitment may depend strongly on the specific study population, intervention, eligibility criteria, and logistical demands of the trial.

However, despite increasing awareness of recruitment difficulties, relatively little is known about how patients are progressively lost throughout the entire recruitment pathway. In particular, previous studies have rarely examined the interaction between the clinical setting in which patients are identified, the stage of post-stroke recovery at which they are screened, and the subsequent logistical requirements of study participation. Studies often report only final enrollment numbers without systematically quantifying attrition at each stage of the recruitment process.

Recruitment may become even more challenging when mechanistic and multimodal approaches are incorporated into neurorehabilitation trials. Such approaches aim to investigate the neural mechanisms underlying recovery and treatment response using neuroimaging and neurophysiological measures in carefully characterized patient cohorts (7). Neuroimaging technologies impose additional technical and patient-related requirements that may further restrict eligibility. While multimodal approaches provide valuable mechanistic insights, their methodological complexity may come at the cost of recruitment feasibility and patient representativeness. This highlights an important trade-off in neurorehabilitation research between obtaining highly controlled mechanistic data and maintaining access to a sufficiently broad and clinically representative patient population.

Therefore, the aim of the present study was to systematically analyze the recruitment process in a stroke neurorehabilitation trial in Germany from initial screening to final enrollment. Specifically, we sought to quantify recruitment efficiency within a real-world clinical rehabilitation setting, identify where patients were lost throughout the recruitment pathway, and determine the relative contribution of clinical, logistical, and study-related barriers to recruitment failure.

By providing a detailed recruitment flow analysis of 1201 screened patients, this study aims to contribute to a better understanding of structural recruitment challenges in stroke neurorehabilitation research and to inform the design of future clinically feasible and more inclusive rehabilitation trials

## 2 Methods

This study was an observational screening analysis conducted at the Rehabilitationszentrum Oldenburg GmbH, Germany. The analysis was embedded within the recruitment process of an exploratory mechanistic pilot study (8) that served as the first step of a larger collaborative research program aiming to identify neural biomarkers of cognitive control as a foundation for future biomarker-informed brain stimulation studies in stroke rehabilitation. The objective of the present study was to systematically evaluate recruitment efficiency and identify barriers to enrollment in a stroke neurorehabilitation study in the German neurorehabilitation setting.

Patient screening took place over a 28-month period between October 2023 to February 2026 in a neurological rehabilitation setting with 60 inpatient beds.

### 2.1 Screening Procedure

All patients admitted to the neurology unit of the rehabilitation center during the study period were screened for potential study participation. A patient was considered “screened” once the medical record had been opened and reviewed by study personnel. Screening was conducted using predefined inclusion and exclusion criteria.

For each patient, the primary reason for exclusion was documented. If multiple exclusion criteria were present, the primary reason for exclusion was determined according to a predefined hierarchical order, with presence of ischemic stroke assessed first, followed by stroke history (single vs. recurrent stroke), followed by age criteria and lastly MRI compatibility. For each patient who was excluded, the first criterion leading to exclusion according to the predefined hierarchical screening sequence was recorded as the primary reason for exclusion. Subsequent exclusion criteria, if present, were not considered in the recruitment flow analysis.

A total of 1201 patients were screened during the recruitment period.

### 2.2 Eligibility Criteria

Inclusion Criteria: Patients were eligible for participation if they fulfilled all of the following criteria:

- history of a single ischemic stroke
- age between 50 and 80 years
- sufficient cognitive and motor abilities to complete study procedures and experimental tasks
- MRI eligibility (i.e., absence of MRI contraindications such as incompatible metallic implants)
- native German language proficiency
- willingness and ability to attend multiple appointments at the university research site

Exclusion Criteria: Patients were excluded if any of the following criteria applied:

- more than one stroke
- history of transient ischemic attack (TIA)
- brainstem involvement
- psychiatric disorders or psychiatric medication
- antiepileptic medication
- severe aphasia or dysarthria
- severe visual neglect
- severe visual impairments, including hemianopia or diplopia
- insufficient cognitive capacity for testing procedures
- Medical comorbidities or medical conditions interfering with participation such as dialysis-dependent chronic kidney disease, dementia or other severe systemic or neurological disorders
- Technical or logistical exclusion criteria
- MRI contraindications
- body dimensions incompatible with MRI head-coil setup
- large distance from the study center (> 100 km)
- inability or unwillingness to repeatedly travel to the university site

### 2.3 Recruitment Flow Analysis

To systematically characterize recruitment barriers, the recruitment process was analysed as a stepwise decision pathway from initial screening to final enrolment. Patients were categorized according to their primary neurological diagnosis, fulfilment of the core study criteria, specific reasons for exclusion, and willingness to participate in the study. Particular attention was given to logistical barriers, especially inability or unwillingness to travel repeatedly to the university-based research site. The overall recruitment process was visualized using a recruitment flowchart and decision-tree model illustrating patient attrition across the different stages of recruitment and eligibility assessment.

### 2.4 Outcome Measures

The primary outcome of the study was recruitment efficiency, defined as the proportion of screened patients who fulfilled the predefined eligibility criteria and the proportion of eligible patients who ultimately consented to participate in the study. Secondary outcomes included the frequency of individual exclusion criteria, the relative contribution of clinical, logistical, and study-related barriers to recruitment failure, and patient attrition across different stages of the recruitment process.

### 2.5 Statistical Analysis

Descriptive statistics were used to summarize screening and recruitment outcomes. Categorical variables are presented as frequencies and percentages. Recruitment pathways and attrition were analyzed descriptively using a decision-tree framework. As the study was exploratory and observational in nature, no inferential statistical analyses were performed.

## 3 Results

Between October 2023 and February 2026, a total of 1201 patients admitted to the neurological rehabilitation center were screened for potential participation in the study. Of all screened patients, 678 patients (56.5%) presented with ischemic stroke and were therefore considered for further eligibility assessment. The remaining patients were excluded at the diagnostic level due to other neurological or non-neurological conditions, including intracerebral hemorrhage (7.9%, n = 95), multiple sclerosis (5.4%, n = 65), tumor resection (5.3%, n = 64), or the absence of central nervous system involvement (e.g. polyneuropathy; 24.9%, n = 299).

Despite the large number of screened patients, only 13 participants ultimately fulfilled all study criteria and agreed to participate in the study. This corresponded to 1.9% of all ischemic stroke patients and 1.1% of the total screened cohort.

The most common reasons for exclusion were clinical in nature. More than half of all ischemic stroke patients (52.2%, n = 354) were excluded due to other clinical exclusion criteria. This category primarily included recurrent stroke events, brainstem involvement, severe comorbidities, psychiatric diagnoses or medication, visual impairments, aphasia or dysarthria, and insufficient German language proficiency. In many cases, patients presented with multiple exclusion criteria simultaneously.

Logistical barriers represented the second largest obstacle to recruitment. A total of 23.9% of ischemic stroke patients (n = 162) were excluded because of travel-related barriers, including excessive travel distance or inability or unwillingness to repeatedly travel to the university study site. Notably, several patients declined participation despite living within the wider regional area surrounding Oldenburg, indicating that even moderate travel demands (<30 km) represented a substantial burden in this clinical population.

Another source of exclusion was the predefined age criterion of 50–80 years, which resulted in the exclusion of 17.9% of ischemic stroke patients (n = 121). Additional exclusion reasons included MRI contraindications (2.2%, n = 15), lack of interest in study participation (1.6%, n = 11), and cognitive impairment preventing completion of study procedures (0.3%, n = 2).

Analysis of the recruitment pathway revealed substantial reduction at multiple stages of the screening process. Initial patient loss occurred at the diagnostic level, as only slightly more than half (56.5%, n = 678) of all screened rehabilitation patients fulfilled the basic criterion of having had an ischemic stroke. Subsequent attrition was mainly driven by strict methodological and clinical requirements, particularly regarding lesion characteristic, comorbidities, psychiatric medication, and MRI compatibility.

In addition to clinical exclusion factors, logistical demands associated with repeated visits to the university-based research facility substantially reduced recruitment efficiency. Overall, the combination of strict eligibility criteria, technical study requirements, and logistical barriers resulted in a highly selective final study population.

**Figure 1:**
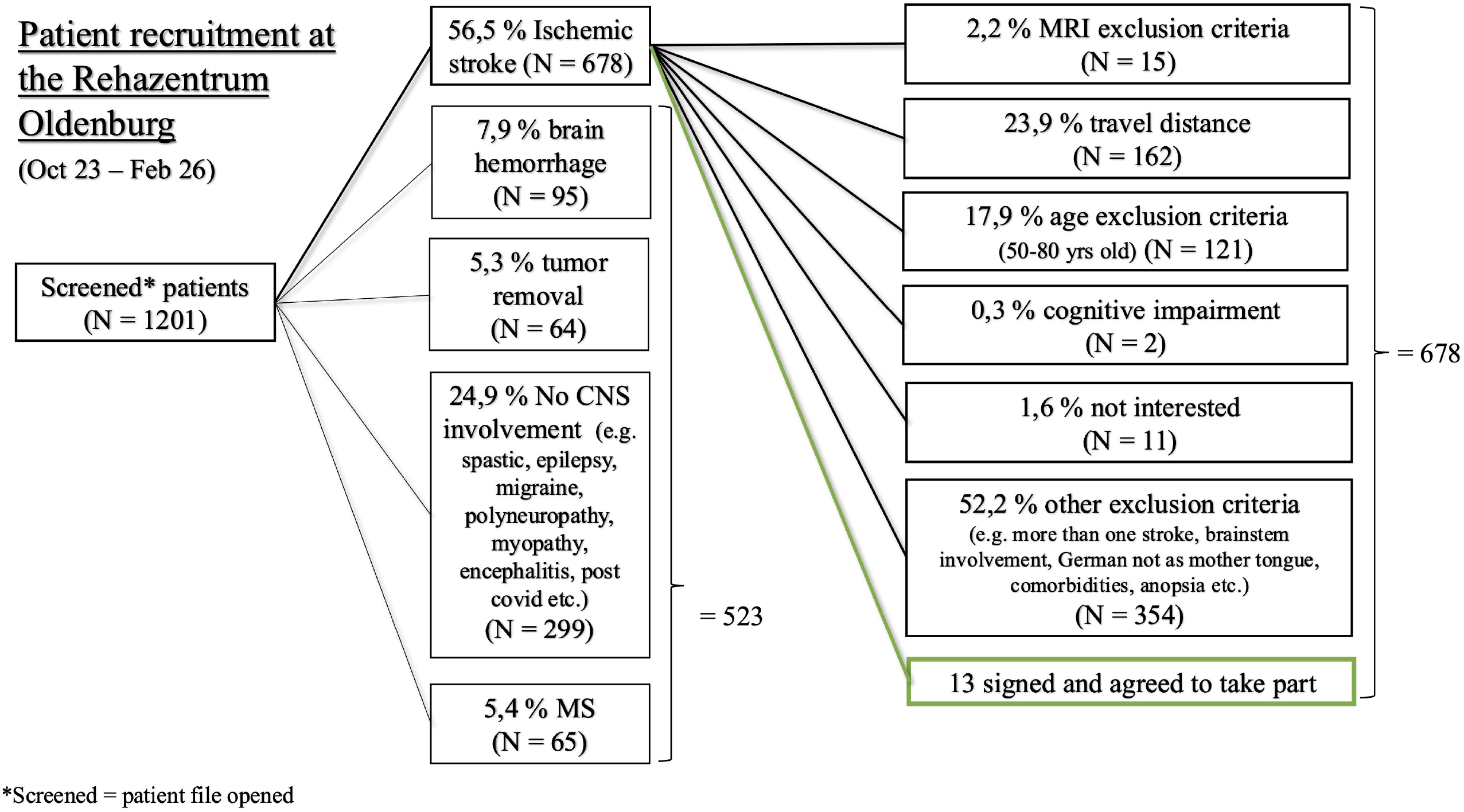
Flow chart of the recruitment process (Frontiers requires figures to be submitted individually)

## 4 Discussion

The present study provides a detailed analysis of recruitment barriers in a real-world stroke neurorehabilitation setting in Germany and demonstrates substantial attrition across multiple stages of the recruitment pathway. Despite screening 1201 patients over a recruitment period of more than two years, only 13 participants were ultimately enrolled. These findings are consistent with previous reports describing extremely low recruitment efficiency in stroke rehabilitation and further extend the literature by systematically quantifying where and why patients are lost throughout the recruitment process.

The largest proportion of patient loss resulted from strict clinical eligibility criteria. In particular, the requirement of a single ischemic stroke substantially reduced the pool of eligible participants. Many otherwise clinically stable and potentially testable patients were excluded due to recurrent stroke events or prior transient ischemic attacks. While restrictive eligibility criteria are commonly implemented to improve cohort homogeneity and internal validity, they may simultaneously reduce external validity, impair recruitment efficiency, and limit the representativeness of study populations (4). Importantly, recurrent and multifocal stroke presentations are highly common in real-world clinical practice (9). Ferrone et al. show that nearly 30% of all strokes are recurrent events. Excluding these patients therefore creates highly selective cohorts that may not adequately reflect the real-world stroke population.

Recent developments within the field appear to increasingly acknowledge this limitation. Contemporary stroke phenotyping studies and multimodal neurorehabilitation projects have started to adopt broader inclusion approaches to improve representativeness and translational relevance. For example, the TiMeS project by Fleury and colleagues aims to establish a multidimensional characterization of stroke phenotypes using multimodal assessments including MRI and transcranial magnetic stimulation (TMS)-EEG in a clinically heterogeneous cohort in a longitudinal study (10). This broader inclusion strategy is particularly noteworthy because highly selective eligibility criteria are often used in mechanistic studies to reduce biological heterogeneity, increase internal validity, and facilitate the interpretation of neurophysiological outcome measures. TiMeS thus illustrates that methodologically demanding multimodal studies can nevertheless be designed to accommodate greater clinical heterogeneity. The present findings underscore the importance of this balance between methodological control and clinical representativeness. Particularly in studies involving MRI, non-invasive brain stimulation, or multimodal biomarker assessments, homogeneous cohorts are often considered necessary to detect mechanistic effects with sufficient statistical sensitivity. However, the present data illustrate that this methodological rigor comes at the cost of substantial patient exclusion and limited external validity. The majority of real-world stroke patients present with complex clinical profiles, including recurrent strokes, comorbidities, variable lesion characteristics, mobility limitations, or cognitive impairments. Consequently, highly controlled study populations may differ substantially from the patient populations encountered in routine clinical rehabilitation. This creates an ongoing tension between conducting “clean” and experimentally controlled studies versus studying clinically representative patients. While highly selective cohorts may facilitate mechanistic insights, overly restrictive recruitment strategies risk producing findings with limited translational relevance for everyday stroke rehabilitation practice.

The predefined age restriction of 50–80 years represented another major source of exclusion in the present study. Nearly one fifth of ischemic stroke patients were excluded solely due to age criteria. Although age restrictions are commonly used to reduce interindividual variability and age-related confounding factors, they may substantially limit recruitment efficiency and reduce generalizability (11). Stroke incidence is increasing both in younger and older populations, and strict upper and lower age thresholds may no longer adequately reflect the demographic reality of stroke care. More flexible age criteria, such as inclusion of all adult patients above 18 years, could therefore improve recruitment while simultaneously enhancing external validity.

In addition to clinical criteria, logistical barriers emerged as a major determinant of recruitment failure. Almost one quarter of ischemic stroke patients were excluded because repeated travel to the university-based study center was not feasible. Notably, even patients living within the regional area surrounding Oldenburg frequently considered travel demands too burdensome. An additional factor contributing to the large and challenging recruitment catchment area may be related to structural changes in the allocation of rehabilitation services within the German healthcare system. In recent years, the German Pension Insurance (Deutsche Rentenversicherung) has increasingly assigned rehabilitation placements nationwide. As a result, many patients admitted to the rehabilitation center originated from geographically distant regions across Germany. Although these patients received inpatient rehabilitation treatment in Oldenburg, participation in the present study required repeated follow-up visits to the university-based research facility after discharge. For many patients, traveling several hundred kilometres for additional study appointments was not feasible. This structural healthcare-related factor likely contributed substantially to the high proportion of patients excluded due to travel distance and highlights how external organizational factors beyond the direct control of study investigators may significantly influence recruitment feasibility in neurorehabilitation research.

This finding highlights that logistical feasibility may represent a critical but often underestimated barrier in neurorehabilitation research, particularly in patient populations with reduced mobility, fatigue, or multimorbidity.

Future studies may therefore benefit from more decentralized and home-based approaches (12) to reduce the logistical burden associated with study participation. The increasing availability of portable neuromodulation systems, tele-neurorehabilitation platforms, and remote cognitive assessments could help reduce transportation-related barriers and improve accessibility. In addition, reimbursement for transportation or taxi services may further facilitate participation in demanding longitudinal study protocols.

The present findings also suggest that some exclusion criteria may have been unnecessarily restrictive. For example, medical comorbidities such as chronic kidney disease or polyneuropathy may not directly interfere with cognitive testing or non-invasive brain stimulation and could therefore be considered more selectively in future trials, with greater emphasis on severe or unstable conditions. Future studies may therefore benefit from differentiating between severe unstable medical conditions and comorbidities that are clinically manageable and unlikely to affect study participation. Similarly, requiring native German language proficiency may unnecessarily exclude otherwise eligible patients. Regarding language-related barriers, future mechanistic stroke studies should increasingly strive to implement language-independent or language-minimized cognitive assessment batteries, thereby reducing language-related exclusion while maintaining the validity of neuropsychological phenotyping. Emerging technology-based approaches, including smartphone-based and largely language-independent assessments, may offer promising opportunities in this regard.

Future neurorehabilitation research may therefore benefit from more pragmatic trial designs that intentionally balance internal validity with external validity and clinical feasibility.

Taken together, the present study supports growing concerns that current neurorehabilitation trial designs may systematically exclude large parts of the real-world stroke population. More pragmatic and clinically inclusive recruitment strategies may therefore be necessary to improve recruitment efficiency, increase representativeness, and enhance the translational value and sample sizes in future stroke rehabilitation research.

## 5 Limitations

Several limitations should be considered when interpreting the present findings. First, the study reflects recruitment procedures from a single rehabilitation center and may therefore not generalize to all clinical settings. Second, exclusion reasons were categorized according to the primary determining factor, although multiple exclusion criteria frequently co-occurred. Consequently, the relative contribution of individual exclusion criteria may partly depend on the hierarchical order in which eligibility criteria were applied. Third, the study focused specifically on a highly demanding neurorehabilitation protocol involving MRI compatibility and repeated visits to a university-based research center, which may have further reduced recruitment rates compared to less intensive study designs.

Despite these limitations, the present study provides a detailed real-world analysis of recruitment attrition across the screening-to-enrollment pathway in stroke neurorehabilitation research. The findings may help inform future trial design and support the development of more feasible, inclusive, and clinically representative rehabilitation studies.

## Data Availability

All data produced in the present study are available upon reasonable request to the authors

## 6 Ethical Considerations

The present study reports on screening and recruitment data collected during the recruitment process of the study by Brückner et al. (2026). The original study protocol was reviewed and approved by the Medical Research Ethics Board of the University of Oldenburg (approval number: 2021–110). The present analysis is limited to data arising from the screening and recruitment process and involved no additional patient contact, assessments, or study procedures.

## 8 Author Contributions

**Anne G. Gerding**: Writing – original draft, Visualization, Investigation, Formal analysis, Data curation. **Christiane M. Thiel:** Writing – review & editing, Supervision, Project administration, Conceptualization, Funding acquisition.

